## Supporting information for "Primary school staff perspectives of school closures due to COVID-19, experiences of schools reopening and recommendations for the future: a qualitative survey in Wales"

**Supplementary File 1: HAPPEN School Staff Survey**

1. What school do you work in?
   a. *(open text response)*
2. What is your role?
   a. *Headteacher*
   b. *Teacher*
   c. *Teaching assistant*
   d. *Support staff*
3. What year do you teach?
   a. *Reception*
   b. *Year 1*
   c. *Year 2*
   d. *Year 3*
   e. *Year 4*
   f. *Year 5*
   g. *Year 6*
4. What does the structure of returning to your school look like?
   a. *Full days*
   b. *Half days*
   c. *Time slots*
   d. *Other*
5. How have you grouped pupils in the classroom?
   a. *By friendship groups*
   b. *By ability*
   c. *By age*
   d. *However we could*
   e. *Other*
6. How many pupils are in each class or group at one time?
   a. *1-30 (select number)*
7. What percentage of your pupils are returning to school before the summer
   a. *0-10%*
   b. *11-20%*
   c. *21-30%*
   d. *31-40%*
   e. *41-50%*
   f. *51-60%*
   g. *61-70%*
   h. *71-80%*
   i. *81-90%*
   j. *91-100%*
8. Has your school involved pupils in its plan to return to school?
   a. *Yes*
   b. *No*
9. If you answered yes, could you provide more detail on how you involved pupils?
   a. *(open text response)*
10. What professional development, training or support do you think you may need as a result of blended learning being implemented for the foreseeable future?
    a. *Teaching and learning approaches*
    b. *Supporting learner health and wellbeing*
    c. *Health and safety*
    d. *Digital skills*
    e. *Other*
11. What other professional development, training or support do you feel would be helpful for returning to school?
    a. *(open text response)*
12. What does your current teaching content include?
    a. *Only teaching the curriculum*
    b. *Teaching some of the curriculum*
    c. *Not teaching any curriculum*
    d. *Other*
13. Since your return to school, how much are you currently delivering teaching outdoors? (outdoor learning)
    a. *Most of the time*
    b. *Some of the time*
    c. *Hardly ever*
    d. *Never*
14. Is this more or less than before lockdown?
    a. *Teaching outdoors more*
    b. *Teaching outdoors less*
    c. *Teaching outdoors the same*
15. How much play time are pupils currently having?
    a. *More than before lockdown*
    b. *The same as before lockdown*
    c. *Less than before lockdown*
16. Do you feel pressure to ensure children are 'catching up' on learning?
    a. *Yes – a lot*
    b. *Yes – a bit*
    c. *No*
17. Please explain your answer above
    a. *(open text response)*
18. Have you seen any benefits of lockdown for children in your class/school?
    a. *(open text response)*
19. Have you seen any negative effects of lockdown for children in your class/school?
    a. *(open text response)*
20. If lockdown happened again, what do you think should be done differently?
    a. *(open text response)*
21. How do you feel about all children returning to school full-time?
    a. *(open text response)*
22. What are your main concerns about returning to school full-time?
    a. *(open text response)*
23. What would help to make this transition easier?
    a. *(open text response)*
24. What should we be evaluating in order to understand the impact of COVID-19 on schools from September?
    a. *(open text response)*
25. Do you have anything else to add about returning to school?
    a. *(open text response)*
